# LEFT ATRIAL APPENDAGE CLOSURE IN PATIENTS WITH MECHANICAL MITRAL VALVE PROSTHESIS: A MULTICENTER ITALIAN PILOT STUDY

**DOI:** 10.1101/2023.12.05.23299544

**Authors:** Alberto Preda, Davide Margonato, Carlo Gaspardone, Vincenzo Rizza, Ciro Vella, Lorenzo Rampa, Alessandra Marzi, Fabrizio Guarracini, Paolo della Bella, Eustachio Agricola, Achille Gaspardone, Matteo Montorfano, Patrizio Mazzone

**Affiliations:** Cardio-Thoraco-Vascular Department, Electrophysiology Unit, ASST Grande Ospedale Metropolitano Niguarda, 20162 Milan, Italy; Cardiovascular Imaging Unit, San Raffaele Scientific Institute, 20132 Milan, Italy; Cardiology Unit, Heart Valve Center, San Raffaele Hospital, 20132 Milan, Italy; Interventional Cardiology Unit, IRCCS San Raffaele Scientific Institute, 20132 Milan, Italy; Department of Cardiac Electrophysiology and Arrhythmology, IRCCS San Raffaele Hospital, 20132 Milan, Italy; Division of Cardiology, San Eugenio Hospital, 00146 Rome, Italy; Interventional Cardiology Unit, IRCCS San Raffaele Scientific Institute, 20132 Milan, Italy; School of Medicine, Vita-Salute San Raffaele University, Milan Italy

**Keywords:** : left atrial appendage closure, mechanical mitral valve prosthesis, atrial fibrillation, stroke, antithrombotic therapy

## Abstract

**Background:** In patients with atrial fibrillation (AF) on vitamin K antagonist (VKA) therapy and therapeutical INR range the incidence of cardiac thromboembolism is not negligible and the subgroup carrying a mechanical prosthetic mitral valve (PMV) has the highest risk. We aimed to assess the long-term effects of left atrial appendage closure (LAAC) in AF patients carriers of mechanical PMV who experienced a failure of VKA therapy.

**Methods:** In this retrospective, multicenter study, patients who underwent LAAC because of thrombotic events including TIA/stroke, systemic embolism and evidence of left atrial appendage thrombosis/sludge during VKA therapy were enrolled. Patients with mechanical PMV were included and compared with controls. The primary endpoint was the composite of all-cause death, major cardiovascular events and major bleedings at follow-up. Feasibility and safety of LAAC was also assessed.

**Results:** A total of 55 patients (42% females; mean age 70 ± 9 years) including 12 carriers of mechanical PMV were enrolled. The most frequent indication to LAAC (71%) was LAA thrombosis or sludge. Procedural success was achieved in 96% of overall cases and in 100% of patients with PMV. In 35 patients a cerebral protection device was used. During a median follow-up of 6.1 ± 4.3 years, 4 patients with PMV and 20 patients without PMV reported adverse events (HR 0.73 [95% CI 0.25 – 2.16, p=0.564]).

**Conclusion:** LAAC seems to be a valuable alternative in AF patients with failure of VKA therapy who are carriers of mechanical PMV. This off-label, real-world clinical practice indication deserve validation in further studies.

## INTRODUCTION

Vitamin K antagonists (VKA) proved to effectively prevent most of cardiac thromboembolism related to AF[1]. However, the residual risk is not negligible and those carrying a mechanical prosthetic mitral valve (PMV) represent the highest risk group (>10% embolic risk per year), despite higher INR targets. Indeed, despite therapeutical INR range and absence of valve thrombosis, the occurrence of cerebrovascular events (CEs) in these patients is not negligible[2,3], approximating 1.7% per year[4]. No studies explored the incidence of possible alternative causes of CEs in patients with AF and PMV neither the incidence of LAA thrombosis in patients on VKA and therapeutical INR range. Moreover, the risk of CEs recurrence in patients on oral anticoagulant (OAC) therapy is reported double compared to patients free from OAC therapy[5] and the presence of LAA sludge/thrombosis is highly predictive of future CEs[6]. Therefore, ischemic stroke and LAA thrombosis during OAC therapy should be equally considered as OAC failure and further prevention strategies are deemed necessary. Clinical guidelines do not provide clear recommendations and therapeutical options in patients with direct oral anticoagulants (DOACs) contraindication are very limited[7]. According to several studies, intensification of OAC was associated with a suboptimal result and a concomitant increased bleeding risk, particularly if INR >3.5[8,9]. Of paramount interest, the last European guidelines on valvular heart disease recommended left atrial appendage closure (LAAC) in all AF patients high thromboembolic risk undergoing valve surgery[10]. This procedure can be performed also by percutaneous approach and is currently the only nonpharmacologic option for preventing CEs in patients with AF at significant stroke risk. LAAC is a continually expanding indication and may play a key role in cases of OAC failure[11], as suggested by newer studies[12], and availability of cerebral protection devices (CPDs) may further improve procedural outcome[13]. However, since patients with LAA thrombosis and mechanical PMV were excluded in all studies, LAAC is currently off-label in this scenario[14]. So far, only a case of LAAC in mechanical PMV and concomitant LAA thrombosis is reported[15]. The aim of this study is to assess the long-term effects of LAAC in AF patients with PMV and failure of VKA therapy as well as the feasibility and safety of the procedure.

## MATERIALS AND METHODS

### STUDY POPULATION

This is a retrospective, multicenter study carried out at the Niguarda Hospital (Milan), San Raffaele Hospital (Milan) and Sant ’Eugenio Hospital (Rome), Italy. Patients were enrolled from January 2012 to January 2022. The following inclusion criteria were considered:

- AF patients on OAC therapy with VKA who underwent percutaneous LAAC;

- Recent history (<1 month prior to LAAC) of failure of appropriate VKA therapy. Events considered were:

- occurrence of transient ischemic attack (TIA), stroke or systemic embolism (SE);

- identification of LAA thrombus or moderate/severe sludge at transesophageal echocardiography (TOE);

As for the thromboembolic events, ischemic stroke was defined as a sudden onset of a focal or global neurological deficit, lasting >24h or <24h but with imaging-documented new or presumed new ischemic lesion; TIA was defined as a neurological dysfunction lasting <24h and without new alteration identified on imaging studies[16]. SE was defined as an abrupt vascular insufficiency associated with clinical or radiological evidence of arterial occlusion in the absence of another likely mechanism. LAA thrombus was considered as the presence of a organized hyperechogenic formation placed in the bottom of LAA. LAA sludge was graded according to universally accepted guidelines in case of an intracavitary echodensity consisting of a prethrombotic state with very pronounced spontaneous echocontrast but without being a thrombus formed[6].

The following exclusion criteria were considered:

- Identification of INR below the therapeutic range in conjunction with the event;

- Possibility of embolic origin different from LAA (PMV thrombosis, left ventricle [LV] apical akinesia, intracardiac shunts and severe carotid stenosis) identified at cardiac imaging.

Baseline clinical characteristics and therapy have been recorded for all patients. Written informed consent was obtained from each patient. The retrospective study was approved by the Ethics Committee of Niguarda Hospital and complies with the Declaration of Helsinki. Data were recorded in a dedicated database in compliance with the ethic committee of our center.

### TECHNICAL ASPECTS OF PROCEDURES

To minimize the risk of complications, intra-procedural TOE monitoring was always conducted. LAAC procedure was performed directly by a senior operator for each center (P.M., M.M. and A.G.) according to universally accepted guidelines[17]. CPDs were used according to the operator’s discretion. Before the release of the device, its position, anchoring and sizing were evaluated. Procedural success was defined as technical success of device deployment without procedure-related major cardiac events: cardiac tamponade, device embolization, periprocedural stroke (within 24 hours from the procedure) or significant peridevice leak (PDL,

≥5 mm at a Nyquist limit of 20–30 cm/s).

### POSTPROCEDURAL MANAGEMENT AND FOLLOW-UP

Information about periprocedural outcomes, including all procedure and device-related adverse events (<7 days) were extracted from patients charts and electronic records. Patients were discharged from the hospital with the aim of maintaining lifelong OAC therapy to prevent the recurrence of ischemic events with a hybrid strategy consisting of LAAC + OAC therapy. For 3 months after discharge, a single antiplatelet drug was added to OAC regimen according to the operator’s discretion. The first clinical follow-up (FU) was performed at 3 months from hospital discharge with a clinical visit + TOE to rule out major complications such as device-related thrombosis (DRT), device embolization, or significant PDL. Additional long-term FU was conducted by clinical visits in our outpatient clinic or, if not available, by telephone interviews with the patient every year. Data concerning medical therapy and adverse events during FU were recorded, in particular death (cardiovascular and all-cause), events (ischemic stroke, TIA, and SE) and major bleedings.

## STUDY OUTCOMES

Patients with PMV were compared with controls. Acute intraprocedural success and complications as well as long term FU have been explored. The primary endpoint was a composite of (1) all-cause death (ACD), (2) major adverse cardiovascular events (MACE) consisting of TIA/ischemic stroke, SE, and (3) major bleeding at FU. Intraprocedural success was defined as correct LAAC device implantation without procedure-related major adverse events.

## STATISTICAL ANALYSES

Categorical variables were expressed as frequencies and percentages and compared with the χ2 or Fisher exact test while continuous variables were expressed as mean [standard deviation (SD)] or median [interquartile range (IQR)] and Student’s t-test and ANOVA test were used as appropriate. Survival and event-free survival were estimated by the Kaplan–Meier method and compared by log-rank test. Analysis was performed by censoring FU at the time of the last FU or at the time of event occurred. A two-tailed p ≤ .05 was considered statistically significant.

Data were analyzed with R version 3.6.2 software (R Foundation for Statistical Computing, Vienna, Austria).

## RESULTS

Overall, 1753 patients who underwent LAAC during the study period were evaluated. LAAC for secondary prevention of OAC failure was performed in 167 patients but only patients who met the above mentioned inclusion criteria were selected. A total of 55 patients (42% females; mean age 70 ± 9 years) were enrolled, of whom twelve (12%) were carriers of mechanical PMV. AF was most commonly permanent (40%), congestive heart failure was present in 22 patients (40%) and a quarter of patients had anamnestic history of major bleeding. All patients were on warfarin therapy. The most frequent indication to LAAC (71%) was LAA thrombosis or sludge while the remaining part was anamnestic history of ischemic cerebrovascular events (ICEs). In proportion, patients with PMV had higher rate of ICEs compared to controls (42% vs. 28%, p=0.36) and the median time from the valve replacement to LAAC was 12.6 ± 5.4 years. No patients had history of SE. Baseline characteristics are presented in Table 1.

**Table 1.**
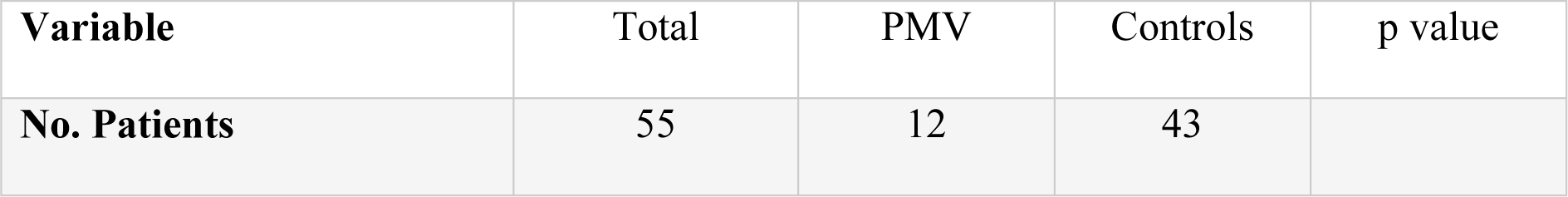
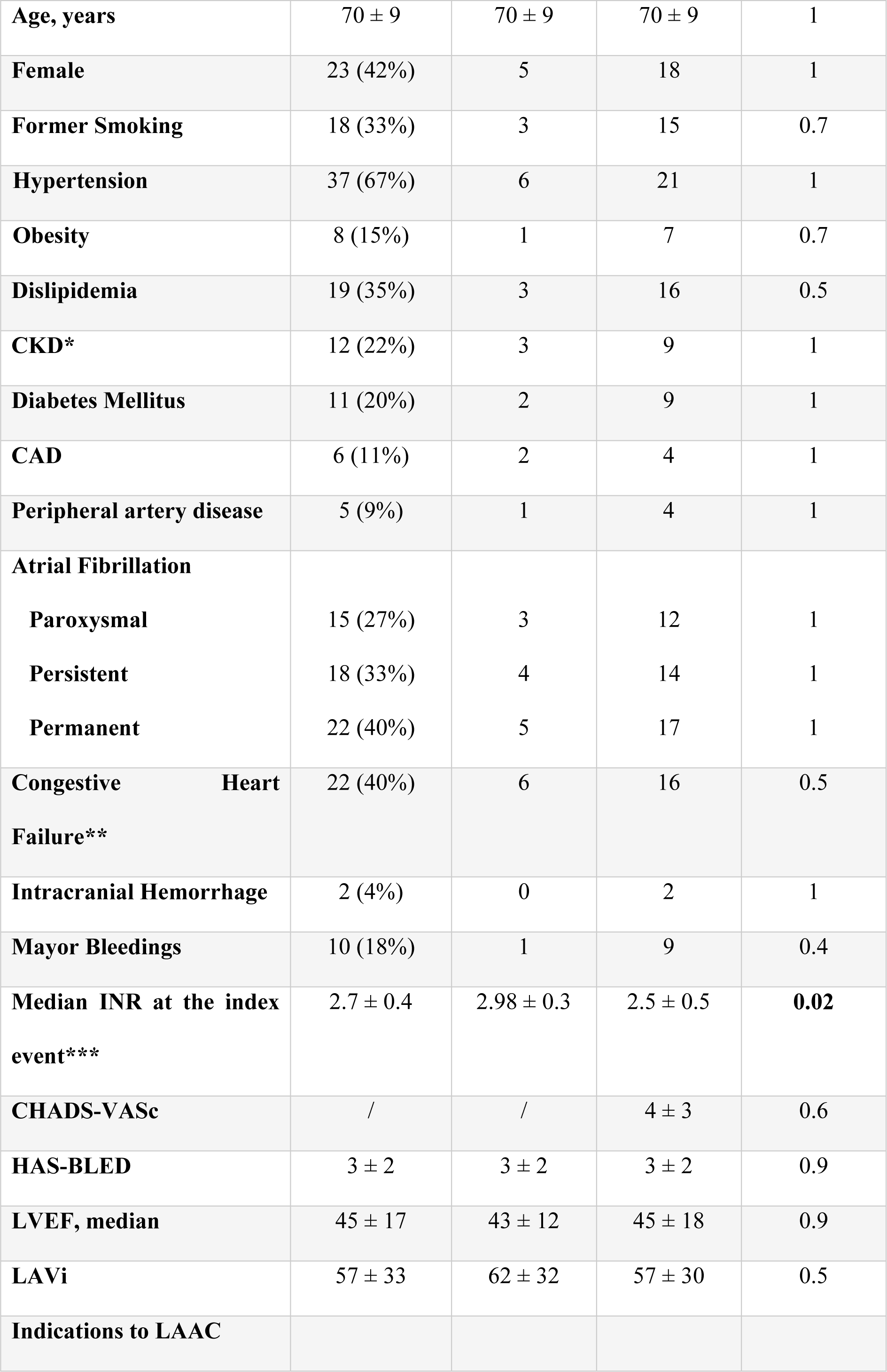
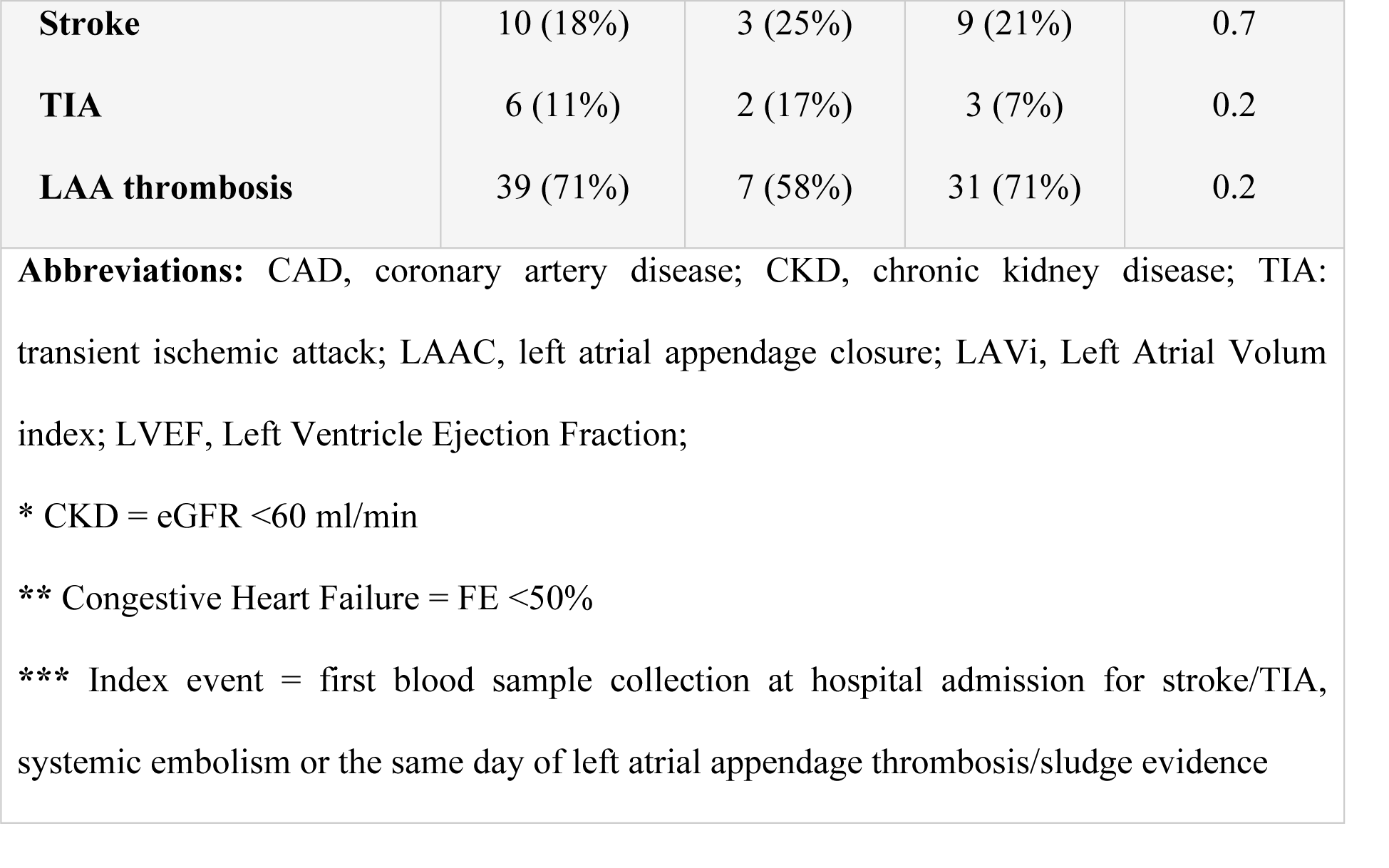
Clinical characteristics of the study population.

| Variable | Total | PMV | Controls | p value |
| --- | --- | --- | --- | --- |
| No. Patients | 55 | 12 | 43 |  |
| **Age, years** | 70 ± 9 | 70 ± 9 | 70 ± 9 | 1 |
| **Female** | 23 (42%) | 5 | 18 | 1 |
| **Former Smoking** | 18 (33%) | 3 | 15 | 0.7 |
| **Hypertension** | 37 (67%) | 6 | 21 | 1 |
| **Obesity** | 8 (15%) | 1 | 7 | 0.7 |
| **Dislipidemia** | 19 (35%) | 3 | 16 | 0.5 |
| **CKD\*** | 12 (22%) | 3 | 9 | 1 |
| **Diabetes Mellitus** | 11 (20%) | 2 | 9 | 1 |
| **CAD** | 6 (11%) | 2 | 4 | 1 |
| **Peripheral artery disease** | 5 (9%) | 1 | 4 | 1 |
| **Atrial Fibrillation** |  |  |  |  |
| **Paroxysmal** | 15 (27%) | 3 | 12 | 1 |
| **Persistent** | 18 (33%) | 4 | 14 | 1 |
| **Permanent** | 22 (40%) | 5 | 17 | 1 |
| **Congestive Heart Failure\*\*** | 22 (40%) | 6 | 16 | 0.5 |
| **Intracranial Hemorrhage** | 2 (4%) | 0 | 2 | 1 |
| **Mayor Bleedings** | 10 (18%) | 1 | 9 | 0.4 |
| **Median INR at the index event\*\*\*** | 2.7 ± 0.4 | 2.98 ± 0.3 | 2.5 ± 0.5 | **0.02** |
| **CHADS-VASc** | / | / | 4 ± 3 | 0.6 |
| **HAS-BLED** | 3 ± 2 | 3 ± 2 | 3 ± 2 | 0.9 |
| **LVEF, median** | 45 ± 17 | 43 ± 12 | 45 ± 18 | 0.9 |
| **LAVi** | 57 ± 33 | 62 ± 32 | 57 ± 30 | 0.5 |
| **Indications to LAAC** |  |  |  |  |
| **Stroke** | 10 (18%) | 3 (25%) | 9 (21%) | 0.7 |
| **TIA** | 6 (11%) | 2 (17%) | 3 (7%) | 0.2 |
| **LAA thrombosis** | 39 (71%) | 7 (58%) | 31 (71%) | 0.2 |

**Abbreviations:** CAD, coronary artery disease; CKD, chronic kidney disease; TIA: transient ischemic attack; LAAC, left atrial appendage closure; LAVi, Left Atrial Volume index; LVEF, Left Ventricle Ejection Fraction;
\* CKD = eGFR <60 ml/min
\*\* Congestive Heart Failure = FE <50%
\*\*\* Index event = first blood sample collection at hospital admission for stroke/TIA, systemic embolism or the same day of left atrial appendage thrombosis/sludge evidence

All patients underwent pre-procedural assessment by TOE to evaluate the LAA size. Overall, 55 devices were successfully implanted and procedural success was achieved in 96% of cases, with one case of cardiac tamponade and one case of PDL. The procedure time was relatively higher in the PMV group (p=0.04), with an average of 12 minutes longer compared to control group. Accordingly, fluoroscopy time was also higher in the PMV group (p=0.02). The most used LAAC device was the Amplatzer Amulet (59%, Abbot Medical) while Watchman, Watchman FLX (Boston Scientific) and Lambre (Lifetech Scientific) were less used. In no cases LAAC device hindered the correct function of the mitral prosthesis. Data about LAAC procedure are summarized in Table 2. In patients with PMV was preferred a diskless device such as Watchman (p=0.07) and Watchman FLX (p=0.02) while in the remaining cases, a deeper implant of the Amulet in the body of the appendage was performed. A CPD between TriGuard3 (Keystone Heart, Caesarea, Israel) and Sentinel (Boston Scientific, Marlborough, MA, US) was positioned in 35 cases judged at high risk of intraprocedural thromboembolism defined as LAAC thrombosis or severe LAA sludge (4+). In cases of moderate-to-severe sludge (3+), CPD was used according to operator’s discretion. After the procedure, 25 (45%) patients added an antiplatelet drug to VKA for 3 months. Cardioaspirin was administered in 18 patients while clopidogrel was chosen in 7 patients. In this time frame, no adverse events were recorded overall. Figure 1 shows periprocedural assessment of LAAC in a patient with mechanical PMV by multimodality imaging.

**Figure 1.**
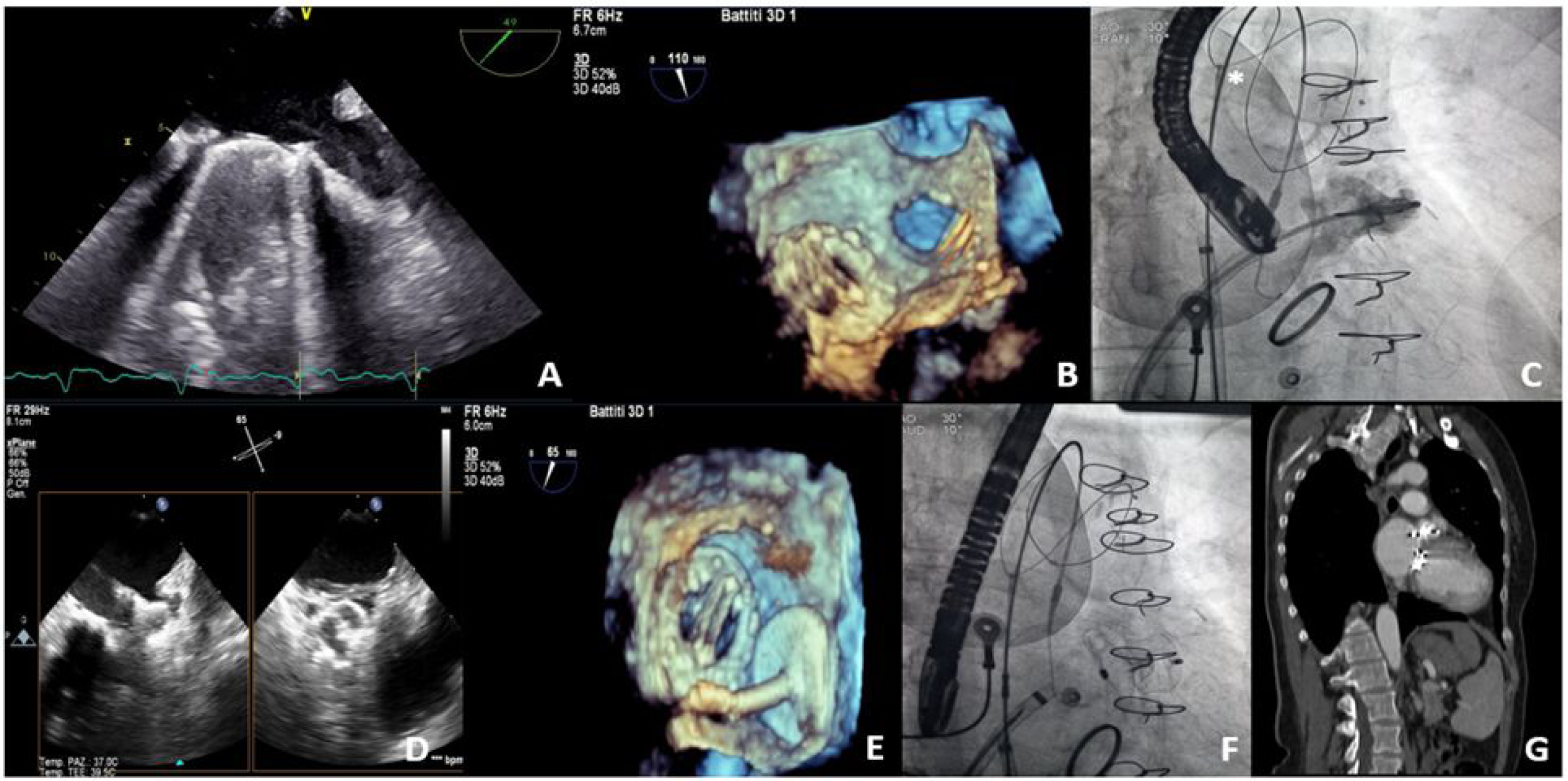
Pre-, intra- and post-procedural assessment of LAAC with multimodality imaging. **Panel A.** *Midesopaheal bicommissural view*: mechanical mitral valve prosthesis and severe spontaneous echo-contrast in LAA; **Panel B**. *3D reconstruction of Midesopaheal bicommissural view*: relationship of mechanical mitral valve prosthesis and LAA orifice into the left atrium. **Panel C**. Fluroscopic view of the injection of the iodine contrast medium into the LAA. The procedure was performed using a cerebral protection device (*); **Panel D**. *Midesophageal MultiView*: positioning of the LAAC device; **Panel E**. *3D reconstruction of Midesopaheal bicommissural view*: relationship between mitral valve prosthesis and the LAAC device; **Panel F**. Fluoroscopic view of the LAAC unhooked from the delivery system; **Panel G**. Post-procedural CT (coronal section) showing mechanical mitral valve prosthesis and LAAC device.

**Table 2.**
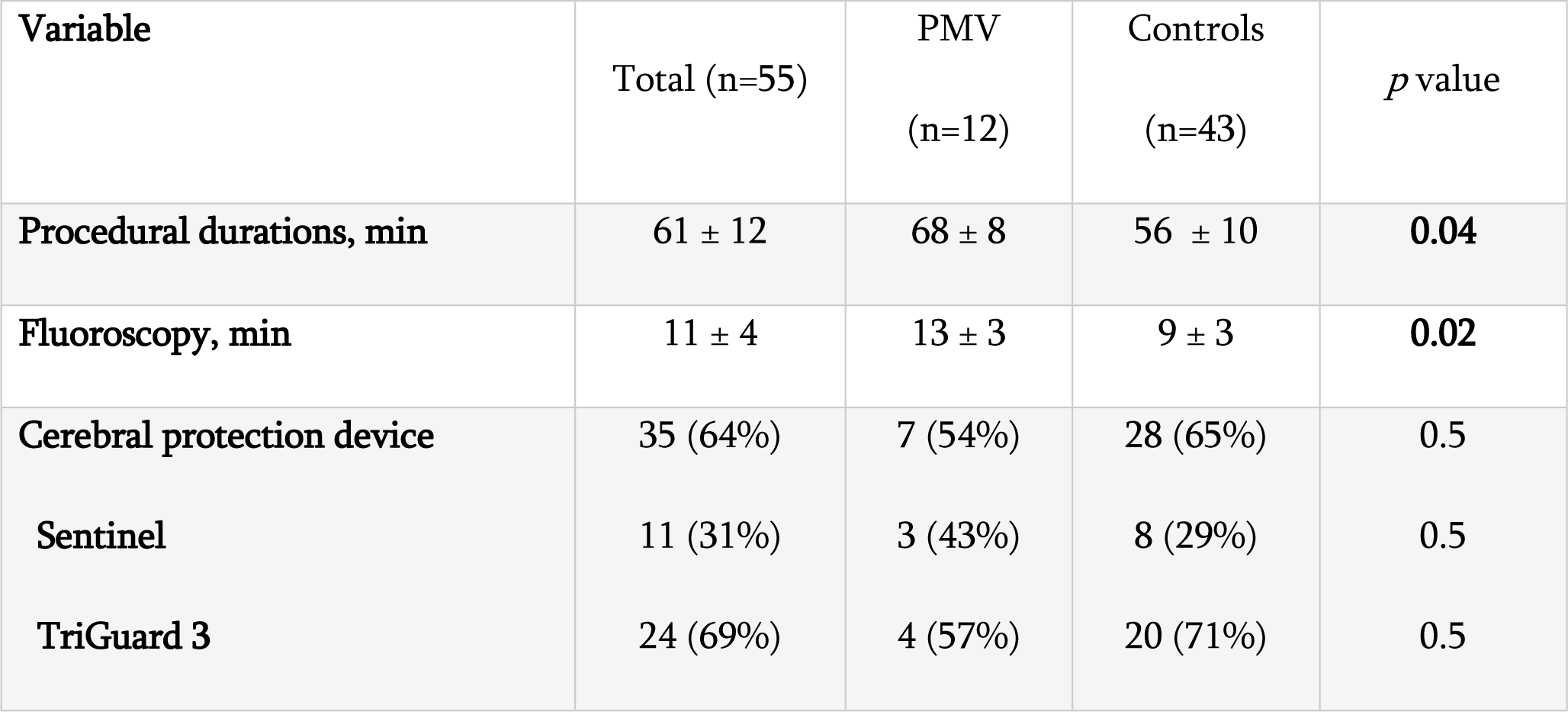
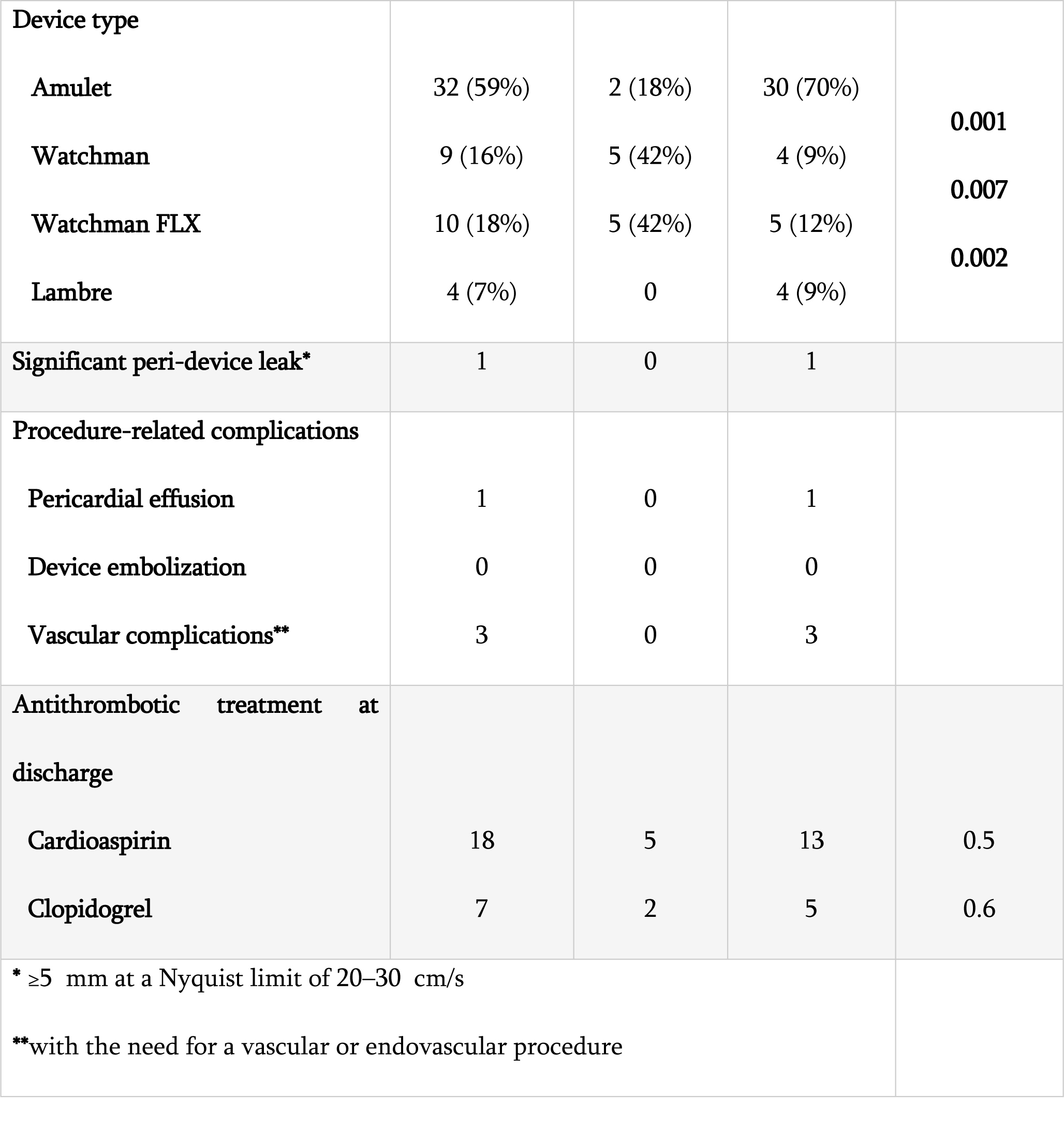
Left atrial appendage closure procedural characteristics.

After discharge from hospital, patients were followed for a median of 6.1 ± 4.3 years. The previous detected peri-device leak was confirmed at the 3-months TOE and no other leaks were noticed. During FU, 9 (16%) patients died for various causes (3 for cardiovascular causes: 2 acute myocardial infarction and 1 pulmonary embolism) of which only 1 from the PMV group (overall median time from LAAC to death 4 ± 5 years). Overall, ICEs occurred in 6 patients (11%) (1 in PMV group versus 5 in control group) while SE occurred in 3 patients (5%) of control group (median time from LAAC to event 3 ± 4 years). Four patients of control group had major bleedings (2 intracranial hemorrhages) that occurred on therapeutical INR range measured at the first day of hospital admission. All the events occurred after discontinuation of the antiplatelet therapy, as reported in Table 3. Kaplan-Meier curves of cumulative survival free of primary endpoint showed no significant differences between groups (Figure 2).

**Figure 2.**
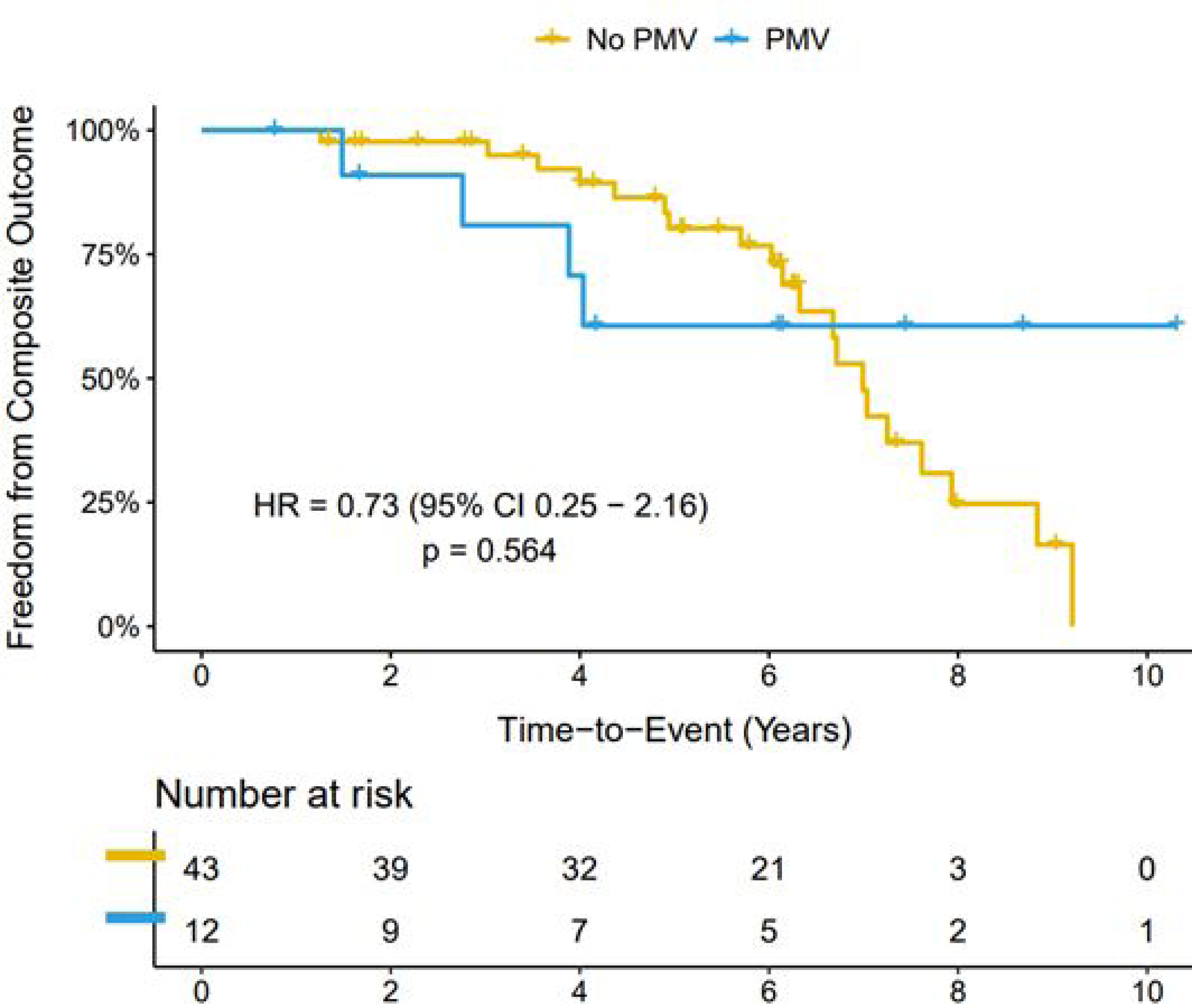
Kaplan-Meier curves of cumulative survival free of primary composite endpoint.

**Table 3.**
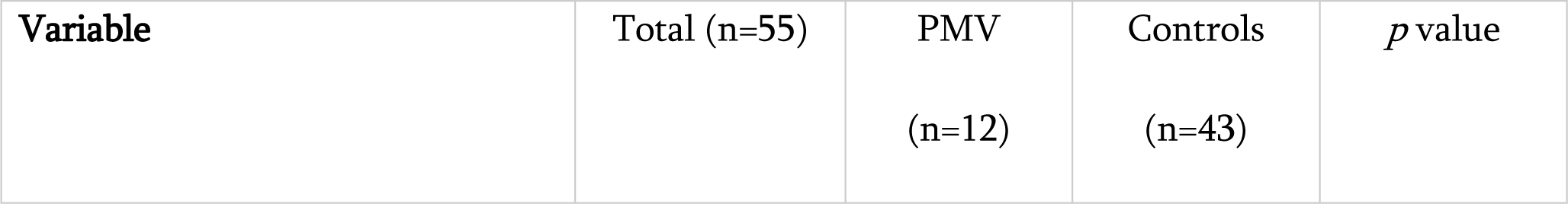
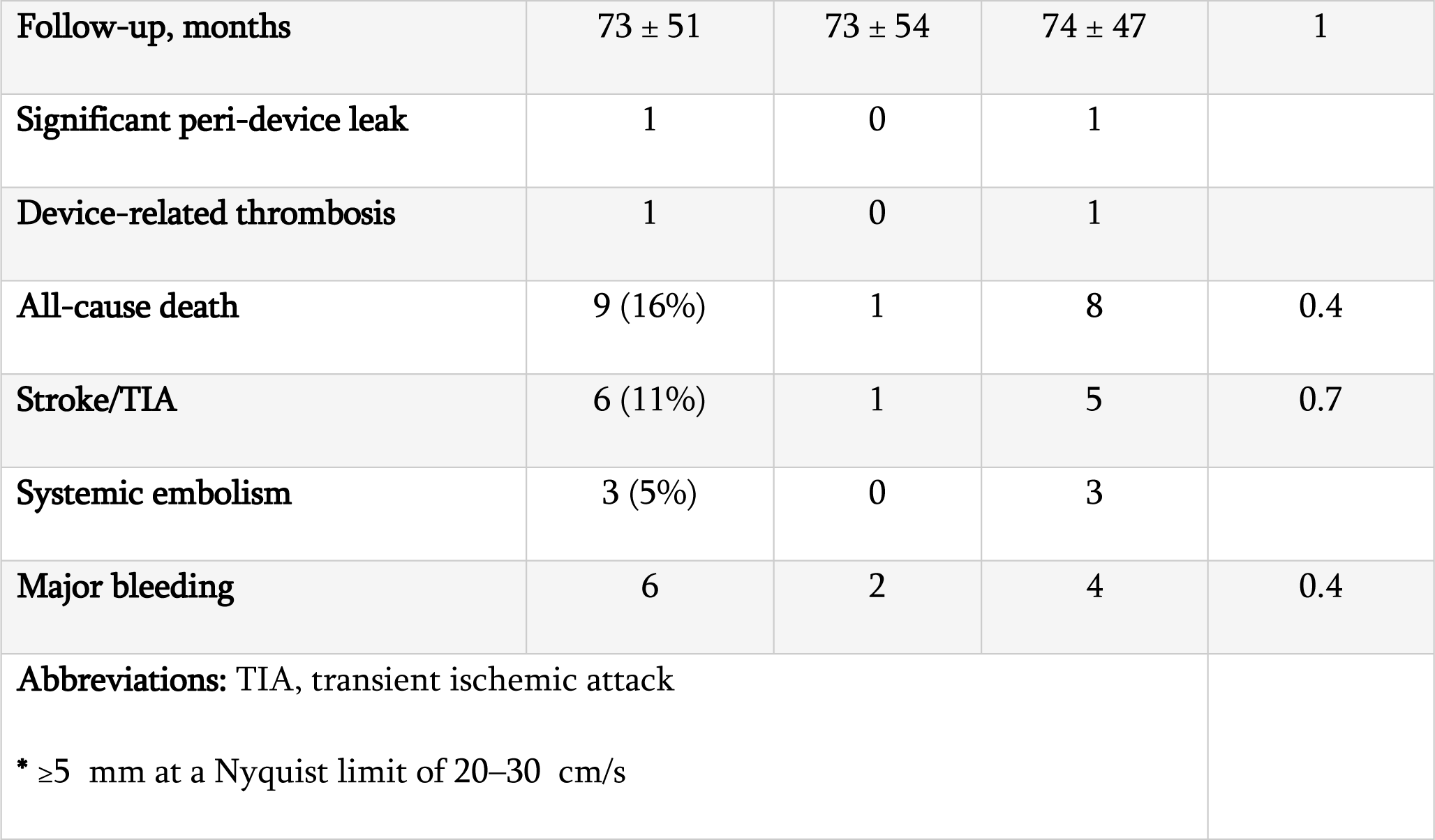
Follow-up and clinical events.

## DISCUSSION

To our knowledge, this is the first study focusing on alternative managements of failure of VKA therapy despite therapeutical INR range in patients with mechanical PMV. By using a control group, we explored both the feasibility and safety and long-term effects of percutaneous LAAC in this specific population. The main findings of this study are: (I) percutaneous LAAC in patients with mechanical PMV is feasible and safe if performed in high volume centers with specialized equips, albeit with longer procedural times; (II) at long-term FU, LAAC in patients with previous PMV is not associated with a higher rate of adverse events compared to control group; (III) no differences were registered in patients who combined antiplatelet therapy with VKA compared to single OAC therapy after the procedure.

### A call for therapeutic alternatives in patients with “valvular AF” and OAC failure

LAAC in patients with AF and failure of OAC therapy is a continually expanding indication due to proved safety and efficacy of the procedure and no need for antithrombotic therapy enhancement[18,19]. However, all LAAC devices that have acquired commercial approvement so far were tested only in the specific population affected by non valvular AF and anticoagulant contraindication. In our study, 167 (9.5%) patients referred to three major Italian centers underwent LAAC due to antithrombotic therapy failure and 12 of them were carriers of mechanical PMV. To be noted, 5 (42%) patients of PMV group had recent history of CE. Since VKAs are the only OAC available in this population, prevention of further adverse events remain a real-world critical concern[20]. LAAC effectiveness in this setting has been confirmed by a recent trial that compared the long-term benefits of LAAC concomitant to cardiac surgery[21]. Enhancement of medical therapy in such scenarios lacks of comparative trials and the decision to improve antithrombotic therapy should be weighted considering several factors such as the bleeding risk[22]. In our study, median HAS-BLED score of the enrolled population was 3, justifying an alternative therapeutic choice.

### Left atrial appendage closure in carriers of mechanical mitral valve prosthesis

AF patients with mechanical PMV are characterized by some additional critical concerns compared to conventional AF patients such as the need for higher INR target and the proximity of the prosthesis to the appendage orifice. In this setting, preprocedural imaging by TOE or cardiac computer tomography are useful to evaluate the relationship between PMV and LAA. Indeed, in cases of little LAA orifice, protruding Coumadin’s ridge or too proximal deposition, the LAAC device may protruding out of the LAA for some millimeters. Despite protrusion of device shoulder by <40% to 50% of its total length is acceptable according to guidelines[23], hindering of valve prosthesis is very unlikely due to the anatomical orientation of LAA and mitral valve. In our study, the major part of PMV group underwent the procedure with an umbrella-shaped device (such as the Watchman), probably due to the higher medical staff experience while in two cases a deeper implant of the Amulet was chosen. Both procedure and fluoroscopy time were higher in PMV group due to the greater complexity and the need to repeated evaluation of the correct positioning of the device on various fluoroscopic and echocardiographic projections. Absence of complications and PDL have been reported in this group. In 35 cases judged at high risk of intraprocedural thromboembolism a TriGuard3 (Keystone Heart, Caesarea, Israel) or Sentinel (Boston Scientific, Marlborough, MA, US) CPD was used. They were constantly employed in cases of LAA thrombosis or severe LAA sludge while in presence of moderate to severe sludge (rating ≤3), a personal choice was performed by the specialist. The use of CPDs in LAAC with LAA thrombosis allowed to a more safe procedure and to reduce the use of specific techniques to minimize interventions within the LAA[24]. So far, a single case report have described a successful deployment of Watchman FLX in the context of LAAC performed with cerebral protection in a patient carrier of mechanical PMV affected by multiple CEs and LAA thrombosis despite therapeutical INR range[15].

### Postprocedural antithrombotic therapy

Our study explored for the first time the effects of a possible hybrid strategy including LAAC plus OAC therapy with VKA in patients with mechanical PMV. All patients continued VKA after LAAC but nearly 50% added an antiplatelet drug for 3 months after LAAC according to the center clinical practice. No DRT were reported and no patients continued antiplatelet therapy beyond that period. Our decision to avoid long-term antiplatelet therapy was in accordance with the only case report present in literature describing a similar scenario[15]. In this case, antithrombotic therapy with VKA alone was continued after the procedure and no CEs during the FU were reported. Currently, the optimal medical therapy after LAAC is still an unresolved issue and varies by type of LAAC device. As this population is highly heterogeneous and characterized by variable thrombotic and bleeding risk, a standard approach is not consistently applied and patients are discharged with different therapeutic regimens according to clinician choice. According to a recent metanalysis, VKA was inferior to DOACs in both CEs and major bleedings endpoints[25]. However, no randomized studies have been performed in patients with indication to continue OAC as well as in patients with mechanical PMV. Since in our specific population the risk of bleeding was not negligible, the hybrid strategy of LAAC plus VKA without the increase of INR target or prolonging antiplatelet therapy was judged the most suitable. At long term FU, despite both higher ischemic and hemorrhagic risk of PMV group compared to control, no differences in terms of adverse events were reported.

### Study limitations

This is an observational, multicenter, retrospective study; therefore, it has the inherent limits of the study design, and our results must be confirmed in a larger sample size. The choice to perform LAAC in patients with mechanical PMV and high-ischemic risk represents an off-label approach based on real-world practice. To compare procedural outcome of LAAC as well as its long term effects, a population with same indication but without mechanical PMV was selected. This choice brings the inevitable bias that PMV group needs higher INR levels than the control group and therefore intrinsically different ischemic and hemorrhagic risk profiles. Finally, so far there are no data available to compare our hybrid approach to an approach of anticoagulant intensification/switch treatment without LAAC: this comparison would indeed be of great importance to evaluate and, eventually, support the role of our strategy.

### Conclusions

In this pilot study we reported the feasibility and safety and the long-term beneficial effects of LAAC in patients with mechanical PMV performed in high volume Italian centers. The present study is a proof of a new concept but further evaluation of this strategy in large prospective, controlled trials is warranted.

## Data Availability

The data that support the findings of this study are available from the corresponding author, Dr. Alberto Preda, upon reasonable request.

## Acknowledgement

none

## Impact on daily practice

Patients with AF and mechanical PMV who had previous failure of VKA therapy despite therapeutical INR range have very limited alternative options. In these patients at very high risk of thromboembolic recurrence, LAAC seems to be a valuable alternative. In our study, LAAC performed in experienced, high volume centers showed low rate of intraprocedural complications and no differences at long-term FU compared with patients without mechanical PMV. The use of CPD might be very useful to reduce intraprocedural cardioembolic events. The present study is a proof of a new concept but further evaluation of this current off-label strategy based on real-world practice in large prospective, controlled trials is warranted.

## Abbreviations and acronyms

ACD: all-cause death
AF: atrial fibrillation
CEs: cerebrovascular events
CPDs: cerebral protection devices
DOACs: direct oral anticoagulants
DRT: device-related thrombosis
FU: follow-up
ICEs: ischemic cerebrovascular events
LAA: left atrial appendage
LAAC: left atrial appendage closure
LV: left ventricle
MACE: major adverse cardiovascular events
OAC: oral anticoagulant
PDL: peridevice leak
PMV: prosthetic mitral valve
SE: systemic embolism
TIA: transient ischemic attack
TOE: transesophageal echocardiography
VKA: Vitamin K antagonists

---

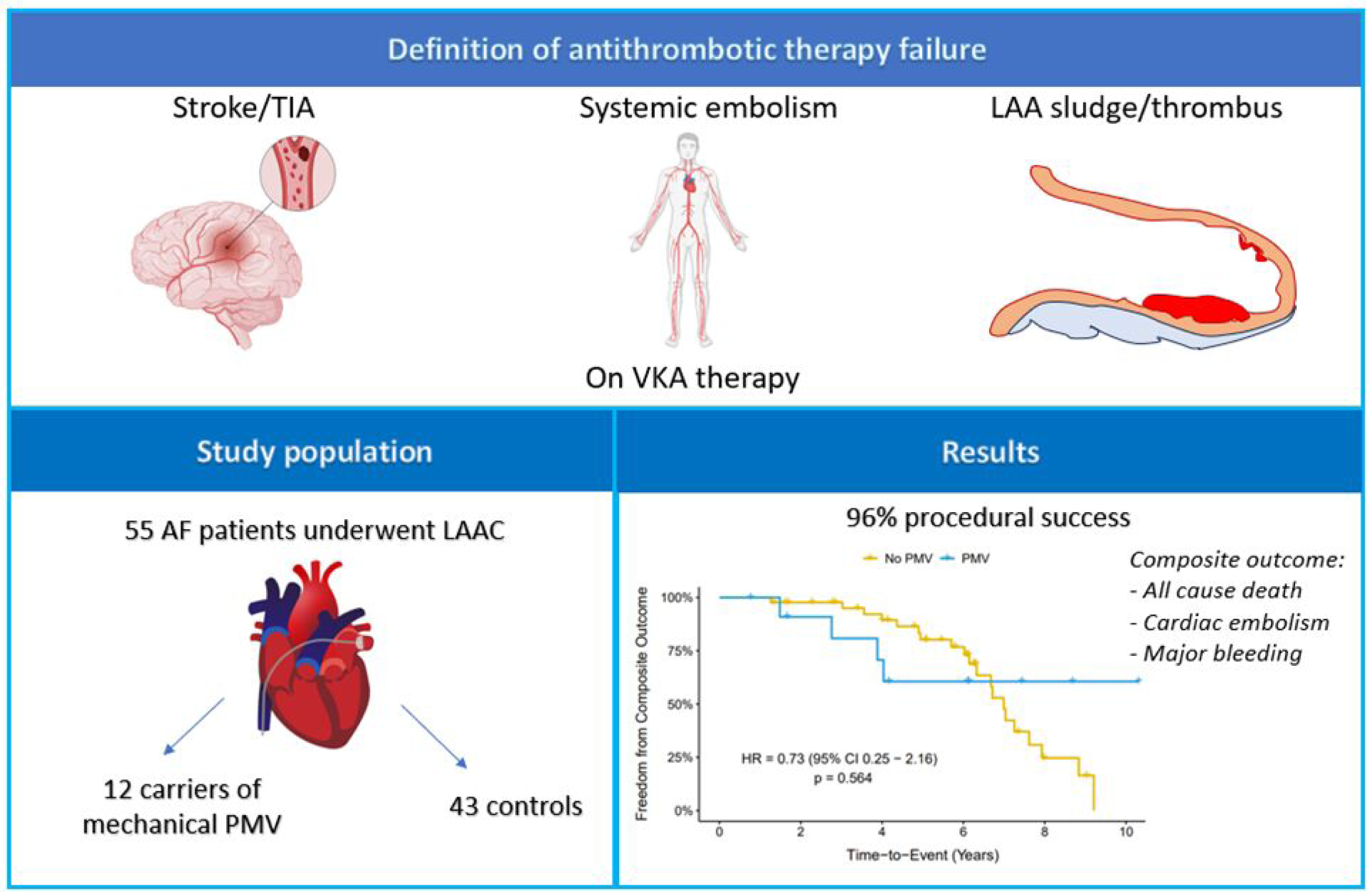
**Central illustration.** Definition of antithrombotic therapy failure. Total enrolled patients who underwent LAAC including those with mechanical prosthetic mitral valve and controls with non valvular AF. Kaplan-Meier survival estimates for the occurrence of the primary endpoint.. LAA, left atrial appendage; LAAC, left atrial appendage occlusion; AF: atrial fibrillation; PMV, prosthetic mitral valve, VKA: vitamin K antagonist

